# Impact of Nativity Status on Dietary Behavior and Obesity Among US Adults

**DOI:** 10.1101/2020.01.06.19015735

**Authors:** Joyce T. Alese, Olatunji B. Alese

## Abstract

**Background/Objectives:** Non-communicable diseases and chronic conditions such as obesity continue to emerge as public health crises in the United States (US) and globally. It is associated with significant morbidity and mortality. We aimed to evaluate how U.S. immigrants compare to native-born adults regarding obesity-related behavior such as dietary intake.

**Subjects/Methods:** The Health Information National Trends Survey was analyzed for this study. The survey was conducted between September and December 2013. Univariable and multivariable logistic regression models were utilized for covariates of interest.

**Results:** 3131 respondents were included in the analysis. Mean age was 54.68 years (SD +/- 16.5) with a female preponderance (61%). Majority were native-born (83%). About 25% of the immigrants were obese, compared to 34% of non-immigrants. After adjusting for gender, age group, race/ethnicity, level of education, marital status and income category, immigrants were more likely to take some quantity of fruit daily (aOR = 1.88; 95% CI: 1.07 - 3.32; p = 0.0290); and less likely to consume soda every week (adjusted OR = 0.74; 95% CI: 0.55 - 0.98; p = 0.0383). Compared to Caucasians, Hispanics (aOR = 2.00; 95% CI: 1.50 - 2.65; p <.0001) and Blacks (aOR = 2.76; 95% CI: 2.08 - 3.64; p <.0001) were more likely to consume soda on a weekly basis.

**Conclusion:** U.S. immigrants are less likely to be obese, and they have healthier dietary behavior compared to non-immigrants. Further studies are needed to determine the effects of various socio-economic, demographic and cultural factors that impact determinants of obesity.

## Introduction

Non-communicable diseases (NCDs) and chronic conditions continue to emerge as public health crises in the United States (U.S.) and globally.^1^ One of the most prominent of such is obesity, a major chronic condition associated with many adverse health outcomes.^2^ Obesity is defined as a body mass index (BMI) equal to or greater than 30kg/m^2^, and more than a third (roughly 35%) of adults in the U.S. is obese.^3^ Individuals’ lifestyle choices have an impact on their BMI, and obesity could be the consequence of a number of lifestyle choices, collectively known as obesity-related behavior.^4-6^

Studies have established the remarkable impact of positive health behavior (including appropriate diet) in reducing obesity rates in general.^7,8^ In view of the steady increase in the number of U.S. immigrants, and the evidence supporting lower risks of obesity among immigrants, it is important to explore and characterize any dissimilarity that may exist in obesity-related behavior, comparing immigrants to native-born U.S. adults. Good knowledge and understanding of any such dissimilarity^9^ will help policy makers and public health practitioners in formulation of policies and tailoring of messages to effectively reduce the risk of obesity in the U.S. populace.

The ‘healthy immigrant’ effect (HIE) posits that new immigrants tend to be healthier than both the native-born populace and immigrants who have lived in the nation for longer periods of time. HIE can be viewed as paradoxical because compared to non-immigrants, many immigrants are of lower socioeconomic status and originate from developing countries. However, several studies have tested and validated the HIE theory.^10-12^ Plausible explanations proffered for HIE in literature include the selective nature of immigration policies of developed countries, including mandatory health and fitness screening requirements.^13^

The current study uses a nationally representative sample of U.S. adults to explore and report proximal determinants of obesity (dietary behavior), comparing U.S. immigrants to non-immigrants. It will add to the research on obesity in the U.S. by providing data that can inform appropriate recommendations for specific and targeted interventions. We hypothesize that adult U.S. immigrants engage in healthier dietary behavior than native-born U.S. adults. Although unlikely to be the sole reasons, healthier dietary behavior would be expected to contribute to the lower risks of obesity observed among immigrants, compared to native-born U.S. adults.

## Materials and Methods

Participants included a nationally representative sample of individuals in the U.S 18 years or older were surveyed in the Health Information National Trends Survey (HINTS). Analysis of the HINTS 4 cycle 3 dataset was conducted under Institutional Review Board (IRB)-exempted protocols. Data Collection was conducted by mail from September 2013 through December 2013 using a protocol similar to that utilized in previous cycles^14^. Data was collected on the American public’s need for, access to, and use of health-related information; as well as data on health-related behaviors, perceptions and knowledge.^15^. The independent variable of main interest in the analyses was the nativity status of participants: immigrant versus native-born. The dependent variables evaluated for the analyses included several indicators of the usual dietary behavior of participants, as well as BMI. For dietary behavior, we selected and examined intake of fruits, vegetables and soda. Covariates of interest were age, gender, level of education, income levels, race/ethnicity, marital and disability status.

### Statistical methods

We performed univariate analyses to describe the distribution of socio-demographic characteristics of survey participants by nativity status. Wald Chi-Square tests (for categorical variables), and t test for differences in means (for the continuous variable ‘age’) were used to determine differences in distributions of demographic variables among immigrants and non-immigrants; and *p*-values <0.05 were taken as statistically significant. Multivariable logistic regression models were used to obtain adjusted odds ratios (AOR) and 95% confidence intervals (95% CI) for associations between nativity status and various outcomes of interest. The various outcomes were taken as indicators of usual dietary behavior among participants and included fruit intake, vegetable intake, soda intake. In all our multivariable models, *p*-values < 0.05 and 95% CI around adjusted odds ratios were used to determine statistical significance. All analyses were done using SAS 9.3 (SAS Institute, Cary NC).

## Results

### Characteristics of respondents

Sixty-one percent of the 3131 participants were female. The mean age in the entire cohort was 54.68 years (SD +/- 16.5). Most participants (34.72%) were in the 50-64year age category, 28.36% were 65 years or older, 23.10% were in the 35-49year age category, and 13.82% were in the 18-34year age category. About 58% of survey participants were White, 18.75% were Hispanic, 15.45% were Black, 4.18% were Asian and 3.49% were of other races. Approximately 52% of respondents were married or living with a partner as married. Also, 52.71% of participants were high school graduates or had attended some college, 37.69% had college degrees or higher and 9.59% had less than high school education. The $10,000 - $49,999 income range was the most predominant in the entire cohort (44.91%), followed by the $50,000 - $99,999 income category (28.48%). Seventeen percent of participants made $100,000 or more per annum, while 9.57% of participants made less than $10,000 per annum. About 66% of participants reported home ownership.

Approximately 17% of participants were immigrants (n = 533), while 83% were native-born (n = 2598) (Table 1). Majority of the immigrants (33.8%) were in the 35-49year age category, while majority of native-born respondents (35.5%) were in the 50-64year age category. Among immigrants, the second most common age group was the 50-64year category (roughly 30.8%), while the 65+ category was the second most common among the native-born, accounting for 30%. The 18-34year age category had the least numbers of respondents among both immigrants and native-born participants. Majority of immigrants (55.3%) were Hispanic, while most native-born respondents (67.3%) were White. The distribution among Non-Hispanic immigrant respondents was as follows: 18.9% Asian, 14.7% White, 9.9% Black and 1.3% other races. Approximately 60% of immigrants, compared to 50% of native-born respondents, were married or living as married.

**Table 1:**
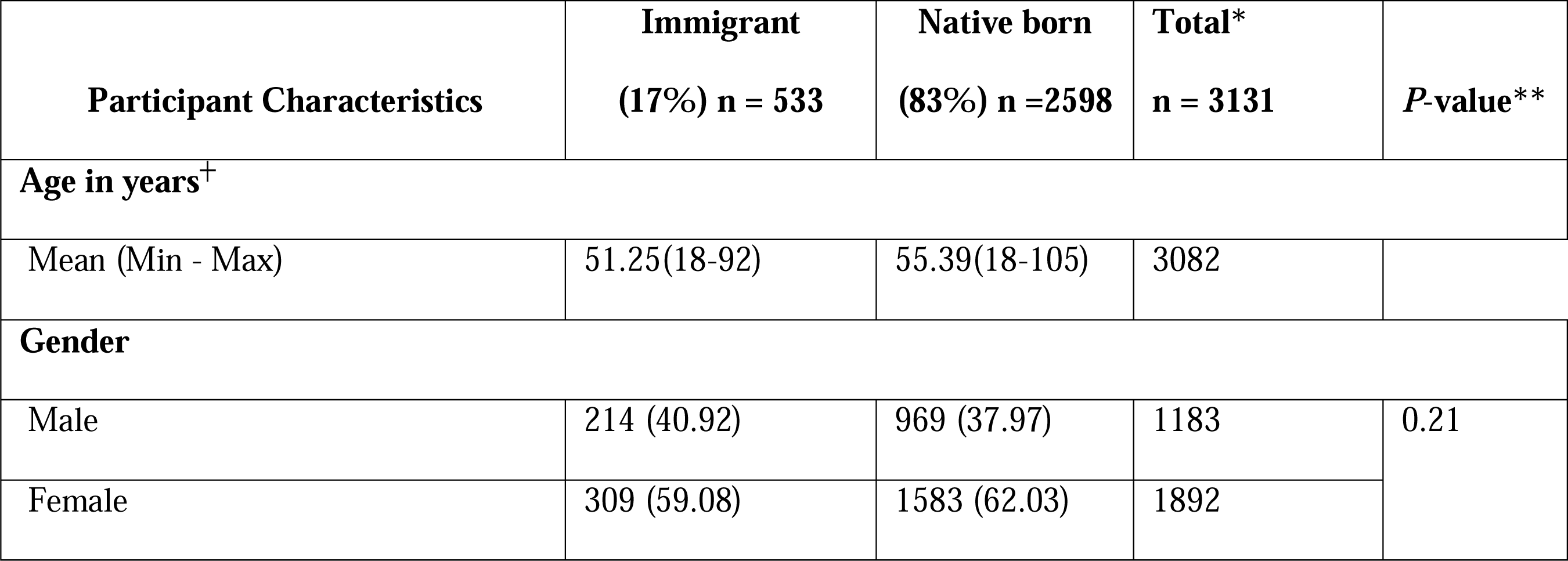
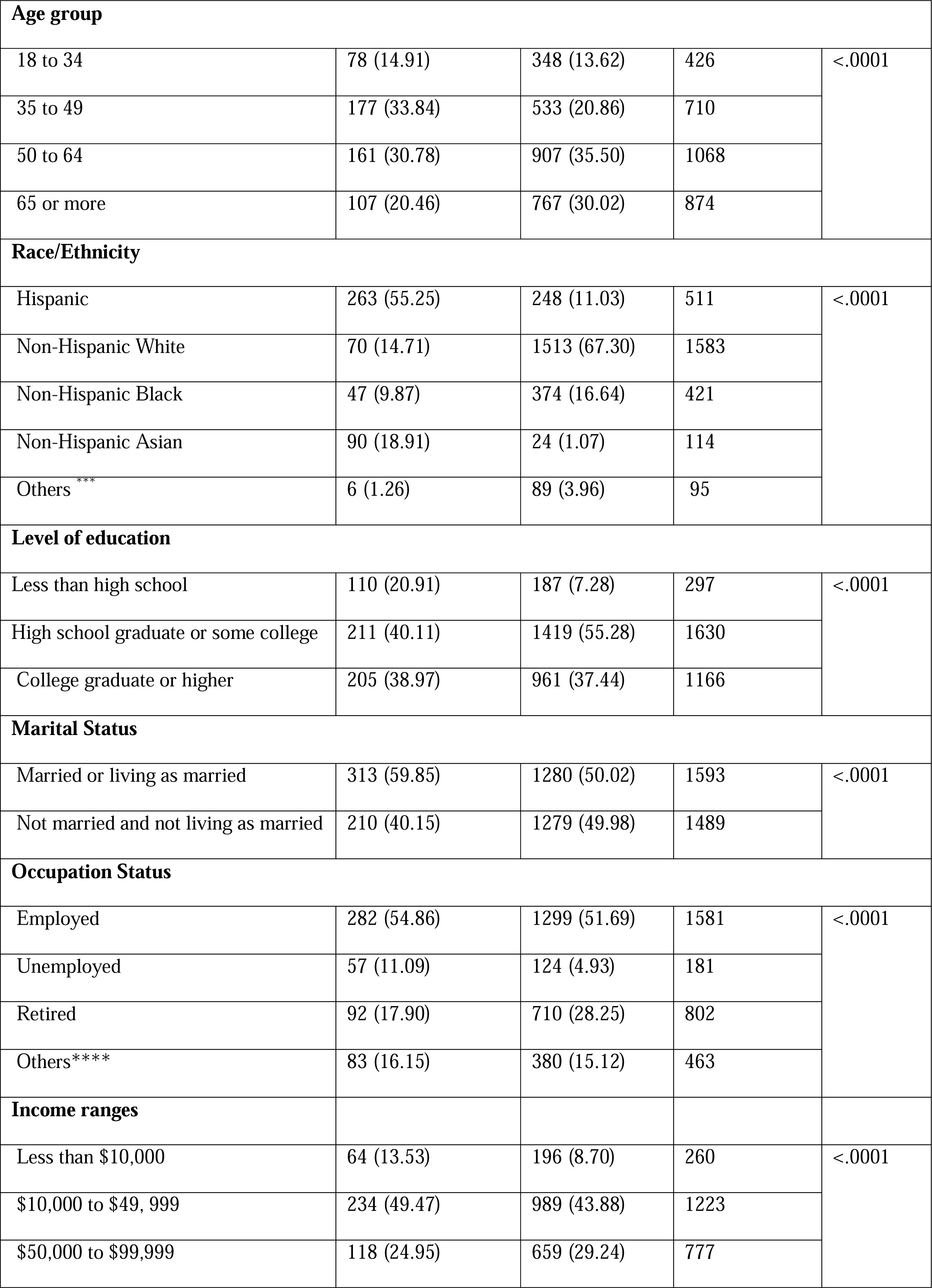
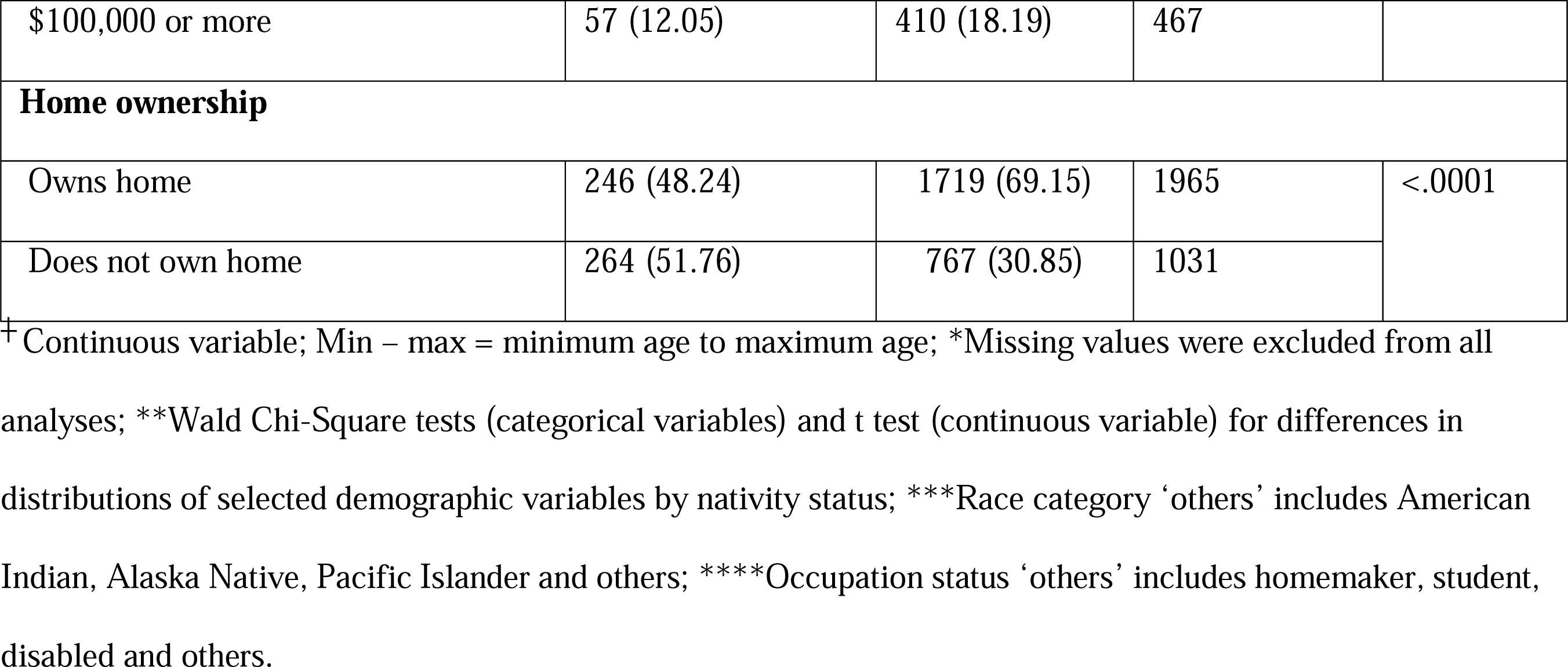
Demographic characteristics by nativity status of participants

Regarding occupational status, 54.9% of immigrants were employed compared to 51.7% of native-born respondents. Eleven percent of immigrants were unemployed, compared to 4.9% of native-born participants. Sixty-three percent of immigrants compared to 52% of native-born respondents had total income less than $50,000 per annum. About 13% of immigrants, compared to 8.7% of non-immigrants earned less than $10,000 per annum. Approximately 12% of immigrants and 18% of non-immigrants earned $100,000 or more per annum. Forty-eight percent of immigrants reported owning their homes compared to 69% of native-born respondents.

### Nativity Status, obesity and Dietary habits

Table 2 shows BMI categories by nativity status, as well as distributions of dietary variables by nativity status of participants. The variables presented were chosen as specific indicators of the dietary behavior of participants, as previously described in methods above. About 33% of participants in the entire cohort were obese and 33.78% was overweight. Most immigrants (36.3%) were in the normal weight category while most native-born respondents (34.4%) were in the obese category. An estimated 25.65% of immigrants were obese compared to 34.4% of native-born respondents.

**Table 2:**
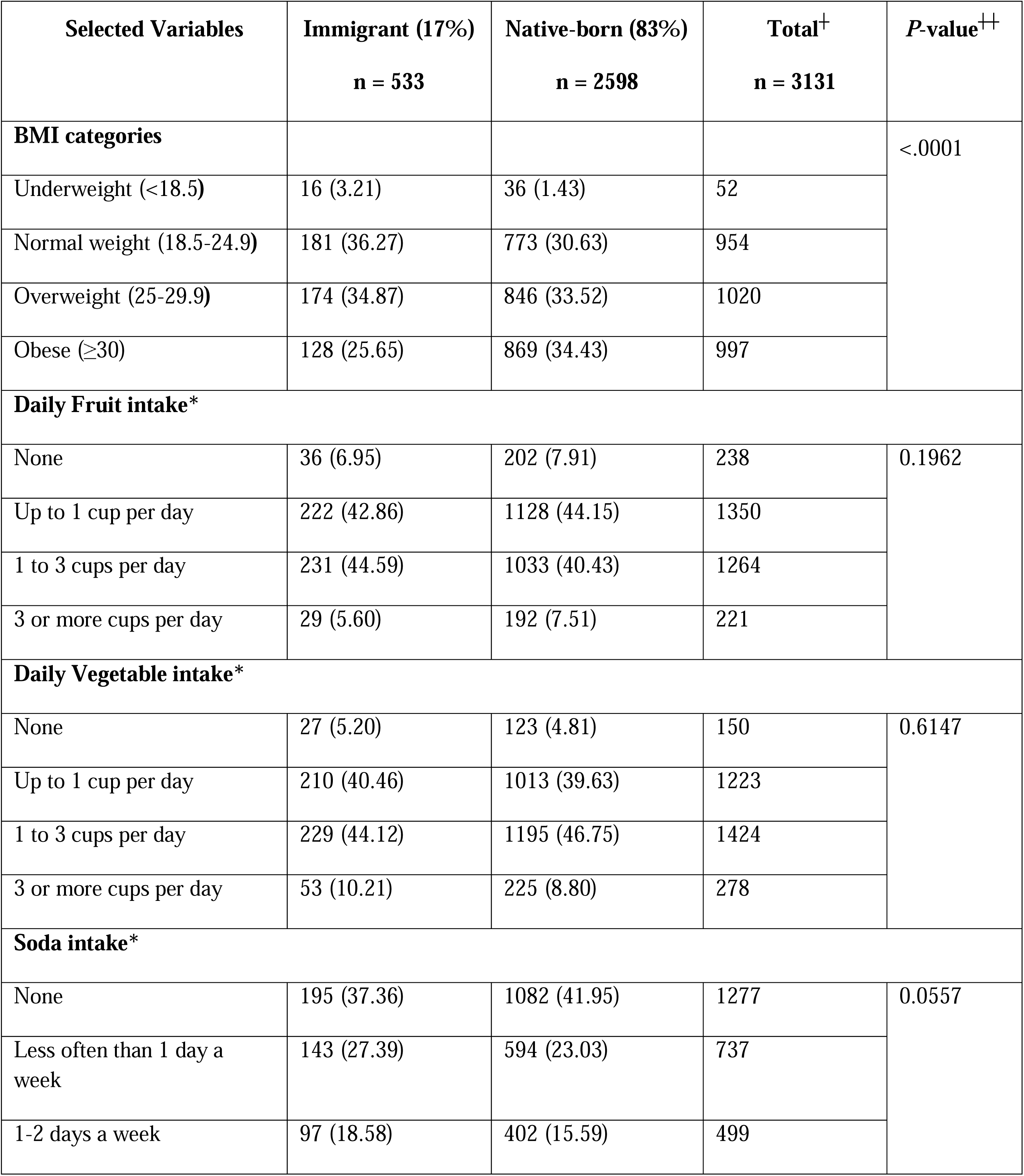
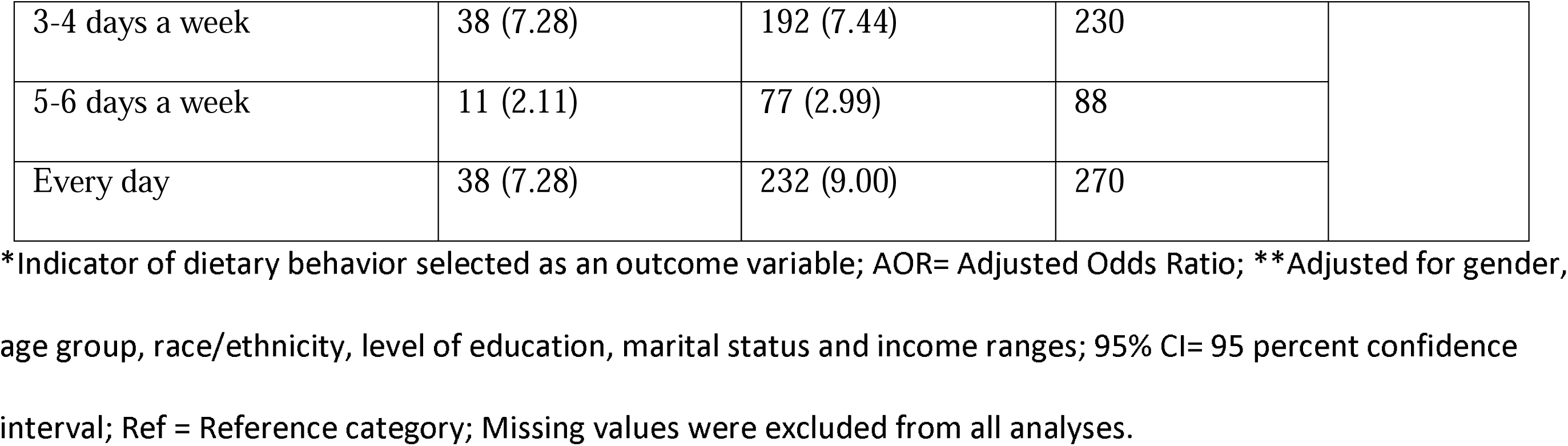
Distributions of indicators of dietary behavior and BMI by nativity status of participants

Most immigrants (44.6%) had a daily fruit intake of 1 to 3 cups per day, while most native-born participants (44.2%) had a daily fruit intake of up to 1 cup per day. Only 5.6% of immigrants and 7.5% of the native-born took 3 or more cups of fruit daily. Approximately 7% of immigrants and 8% of the native-born did not take any quantity of fruit daily. For vegetable intake, most participants took 1 to 3 cups per day: 44.1% of immigrants vs. 46.8% of the native-born. About 5% of immigrants and 4.8% of the native-born did not take any quantity of vegetables daily. About 16% of immigrants and 19% of non-immigrants reported taking soda on 3 or more days per week, while 7.3% of immigrants and 9% of the native-born reported taking soda every day. Thirty-seven percent of immigrants and 41.9% of the native-born reported not taking soda at all.

### Nativity Status and Co-variates of interest

Three separate multivariable logistic regression analyses were performed to estimate associations between nativity status and fruit intake, nativity status and vegetable intake, as well as nativity status and soda intake (Table 3). After adjusting for gender, age group, race/ethnicity, level of education, marital status and income category, immigrants were more likely than native-born respondents to take some quantity of fruits daily (Adjusted OR = 1.88; 95% CI: 1.07 - 3.32; *p* = 0.0290). Although immigrants were also more likely than native-born participants to take some vegetables daily, the association was not statistically significant (Adjusted OR = 1.29; 95% CI: 0.68 - 2.47; *p* = 0.4372). Furthermore, immigrants were less likely than native-born respondents to consume soda every week (Adjusted OR = 0.74; 95% CI: 0.55 - 0.98; *p* = 0.0383).

**Table 3:**
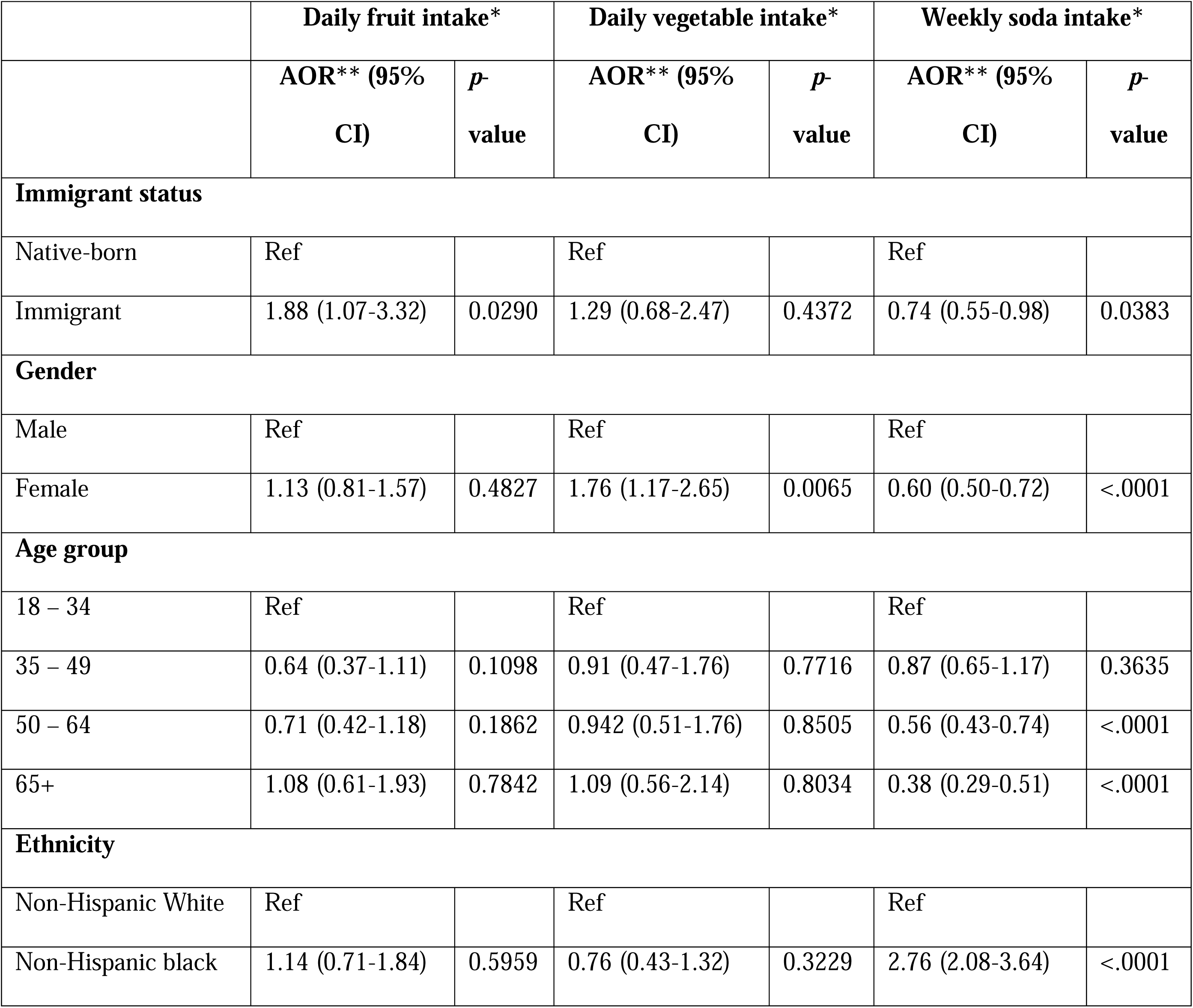
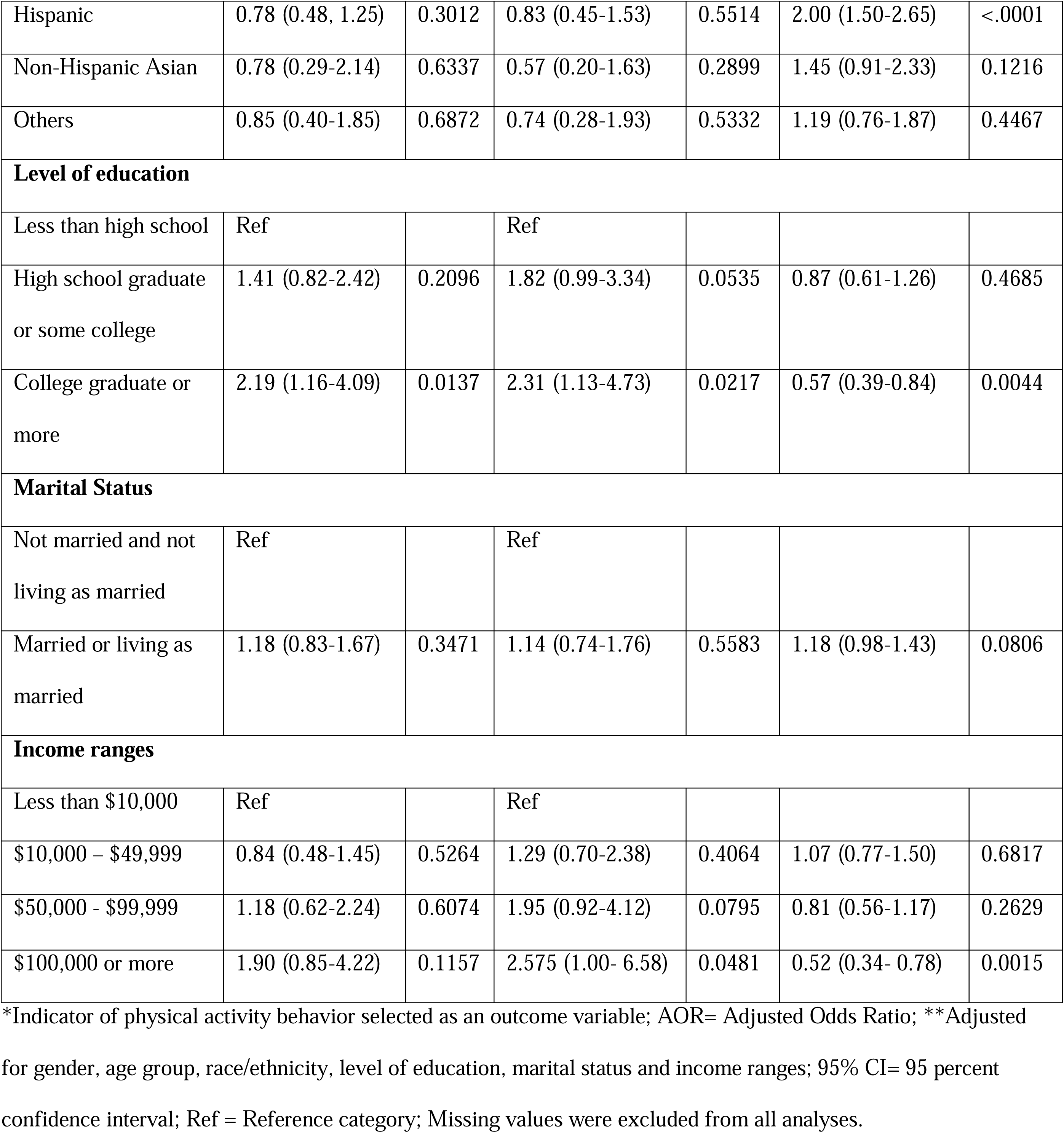
Adjusted logistic regression analyses for associations between nativity status and three separate indicators of dietary behavior among participants.

|  | Daily fruit intake* |  | Daily vegetable intake* |  | Weekly soda intake* |  |
| --- | --- | --- | --- | --- | --- | --- |
|  | AOR** (95% CI) | *p*-value | AOR** (95% CI) | *p*-value | AOR** (95% CI) | *p*-value |
| **Immigrant status** |  |  |  |  |  |  |
| Native-born | Ref |  | Ref |  | Ref |  |
| Immigrant | 1.88 (1.07-3.32) | 0.0290 | 1.29 (0.68-2.47) | 0.4372 | 0.74 (0.55-0.98) | 0.0383 |
| **Gender** |  |  |  |  |  |  |
| Male | Ref |  | Ref |  | Ref |  |
| Female | 1.13 (0.81-1.57) | 0.4827 | 1.76 (1.17-2.65) | 0.0065 | 0.60 (0.50-0.72) | <.0001 |
| **Age group** |  |  |  |  |  |  |
| 18 – 34 | Ref |  | Ref |  | Ref |  |
| 35 – 49 | 0.64 (0.37-1.11) | 0.1098 | 0.91 (0.47-1.76) | 0.7716 | 0.87 (0.65-1.17) | 0.3635 |
| 50 – 64 | 0.71 (0.42-1.18) | 0.1862 | 0.942 (0.51-1.76) | 0.8505 | 0.56 (0.43-0.74) | <.0001 |
| 65+ | 1.08 (0.61-1.93) | 0.7842 | 1.09 (0.56-2.14) | 0.8034 | 0.38 (0.29-0.51) | <.0001 |
| **Ethnicity** |  |  |  |  |  |  |
| Non-Hispanic White | Ref |  | Ref |  | Ref |  |
| Non-Hispanic black | 1.14 (0.71-1.84) | 0.5959 | 0.76 (0.43-1.32) | 0.3229 | 2.76 (2.08-3.64) | <.0001 |
| Hispanic | 0.78 (0.48, 1.25) | 0.3012 | 0.83 (0.45-1.53) | 0.5514 | 2.00 (1.50-2.65) | <.0001 |
| Non-Hispanic Asian | 0.78 (0.29-2.14) | 0.6337 | 0.57 (0.20-1.63) | 0.2899 | 1.45 (0.91-2.33) | 0.1216 |
| Others | 0.85 (0.40-1.85) | 0.6872 | 0.74 (0.28-1.93) | 0.5332 | 1.19 (0.76-1.87) | 0.4467 |
| **Level of education** |  |  |  |  |  |  |
| Less than high school | Ref |  | Ref |  |  |  |
| High school graduate or some college | 1.41 (0.82-2.42) | 0.2096 | 1.82 (0.99-3.34) | 0.0535 | 0.87 (0.61-1.26) | 0.4685 |
| College graduate or more | 2.19 (1.16-4.09) | 0.0137 | 2.31 (1.13-4.73) | 0.0217 | 0.57 (0.39-0.84) | 0.0044 |
| **Marital Status** |  |  |  |  |  |  |
| Not married and not living as married | Ref |  | Ref |  |  |  |
| Married or living as married | 1.18 (0.83-1.67) | 0.3471 | 1.14 (0.74-1.76) | 0.5583 | 1.18 (0.98-1.43) | 0.0806 |
| **Income ranges** |  |  |  |  |  |  |
| Less than \$10,000 | Ref | | Ref | | | |
| \$10,000 – \$49,999 | 0.84 (0.48-1.45) | 0.5264 | 1.29 (0.70-2.38) | 0.4064 | 1.07 (0.77-1.50) | 0.6817 |
| \$50,000 - \$99,999 | 1.18 (0.62-2.24) | 0.6074 | 1.95 (0.92-4.12) | 0.0795 | 0.81 (0.56-1.17) | 0.2629 |
| \$100,000 or more | 1.90 (0.85-4.22) | 0.1157 | 2.575 (1.00- 6.58) | 0.0481 | 0.52 (0.34- 0.78) | 0.0015 |

\*Indicator of physical activity behavior selected as an outcome variable; AOR= Adjusted Odds Ratio; \*\*Adjusted for gender, age group, race/ethnicity, level of education, marital status and income ranges; 95% CI= 95 percent confidence interval; Ref = Reference category; Missing values were excluded from all analyses.

Our multivariable analyses also revealed that female participants were more likely than males to take some quantity of vegetables daily (Adjusted OR = 1.76; 95% CI: 1.17 - 2.65; *p* = 0.0065). We found no significant difference in consumption of fruits and vegetables by age category but respondents with at least a college degree were more likely to have some daily intake of fruits (Adjusted OR = 2.19; 95% CI: 1.16 - 4.09; *p* = 0.0137) and some daily intake of vegetables (Adjusted OR = 2.31; 95% CI: 1.13 - 4.73; *p* = 0.0217). There was no statistically significant difference in daily fruit consumption by income category but respondents who earned $100,000 or above were more likely to take vegetables daily (Adjusted OR = 2.575; 95% CI: 1.00 - 6.58; *p* = 0.0481)

Females were less likely than males to take soda every week (Adjusted OR = 0.60; 95% CI: 0.50 - 0.72; *p* <.0001). Also, older participants were less likely to take soda every week, and this finding was especially notable when comparing the 50 – 64year age category (Adjusted OR = 0.56; 95% CI: 0.43 - 0.74; *p* <.0001) and the 65+ age category (Adjusted OR = 0.38; 95% CI:0.29 - 0.51; *p* <.0001) to the 18 – 34year age category. Respondents with at least a college degree were less likely to drink soda every week (Adjusted OR = 0.57; 95% CI: 0.39 - 0.84; *p* = 0.0044). Respondents with an annual income of $100,000 or above were also less likely to drink soda every week (Adjusted OR = 0.52; 95% CI: 0.34 - 0.78; *p* = 0.0015). Compared to Caucasians, Hispanics (Adjusted OR = 2.00; 95% CI: 1.50 - 2.65; *p* <.0001) and Blacks (Adjusted OR = 2.76; 95% CI: 2.08 - 3.64; *p* <.0001) were more likely to consume soda on a weekly basis.

## Discussion

One of the aims of the current thesis was to describe the demographics of the survey participants by nativity status. A continued look at the demographics of immigrants will produce a better understanding of the impact of recent immigration and serve as a strategy for appraisal of policies related to immigrants and immigration.

Interestingly, our analysis revealed that a much greater proportion of immigrants had less than high school diploma compared to the proportion among non-immigrants. This finding has also been previously documented in the 2011 Current Population Survey (CPS) which showed roughly 28% of immigrants aged 25 to 65 years (compared to an estimated 7% of non-immigrants) without high school certificate.^16^ Level of education is a strong predictor of socio-economic class. It is therefore not surprising to have observed in our analysis that compared to their native-born counterparts, a greater proportion of the immigrant participants were of lower socio-economic status: a higher percentage of immigrants were unemployed, and a lower percentage reported home ownership.

The higher proportion of underweight observed among immigrant participants could reflect the heterogeneity of country of origin; many immigrants originate from developing countries with high poverty levels and food scarcity due to famine. Our bivariate analyses showed that proportions for indicators of diet were similar or close between the 2 groups of interest: some daily fruit intake (93.05% among immigrants vs. 92.09% among non-immigrants), some daily vegetable intake (94.79% among immigrants vs. 95.18% among non-immigrants), and some weekly soda intake (7.28% among immigrants vs. 9% among non-immigrants).However, following adjustments in multivariable analyses, immigrants were significantly more likely than non-immigrants to take fruits daily and significantly less likely to take soda every week. This trend follows current recommendations for healthy living by various advocates.

Notwithstanding the finding that immigrants in the survey engage in healthier dietary behavior than native-born respondents, it is noteworthy that up to 7% and 5% of participants have no daily intake of fruits and no daily intake of vegetables respectively. Furthermore, 16 - 19% of participants consume soda on 3 or more days per week. A prior analysis showed that intake of fruits and vegetables is critical to promotion of good health and that diets rich in fruits and vegetables reduce the risk of obesity, cancer and other chronic diseases.^17^ Furthermore, soda is one of the major sources of added sugars in the diet of the American populace and excess sugar intake has been linked to numerous metabolic problems, adverse health outcomes and deficits of essential nutrients.

The American Heart Association (AHA) recommends that notwithstanding intake of diets rich in fruits and vegetables, minimizing intake of beverages and foods with added sugars is necessary for healthy living.^18^ Researchers, practitioners and policy makers need to develop targeted strategies and focus attention on keeping immigrants in the loop of positive health behavior, encouraging older adults to engage in more physical activity, and increasing the level of education and the earning power of the general public.

To our knowledge, this is the first detailed review of dietary behavior between immigrants and native-born U.S. adults. The HINTS dataset has been validated to capture a representative proportion of U.S. adults regarding their health practices including dietary behavior. Limitations of our study include retrospective analysis with inherent potential biases and confounders. In addition, social and personal factors of the participants that influenced the choice of specific health and dietary behaviors cannot be ascertained from the analysis of this dataset. However, this study showed major differences in dietary behavior between U.S. immigrants and non-immigrants and serves as a template on which more robust studies on obesity-related behavior can be built.

Further studies specific to participants from specific ethnic, racial and nationality groups would enable a more robust mechanism for ascertaining the impact of demographic characteristics on obesity risks. Future research embracing these types of studies would be useful in expanding knowledge and understanding of elements that may differentiate U.S. immigrants from native born from adults in their propensity for developing obesity.

In conclusion, we reported an association of higher healthy dietary lifestyle and behavioral choices with lower proportions of obesity among immigrants. In the general cohort, increase in age was significantly associated with increase in obesity-related behavior, while higher levels of education and higher income were associated with decrease in obesity-related behavior. Previous studies have shown that notwithstanding obesity risk status, most individuals will derive benefits from better nutrition and healthy eating choices. Public health researchers and practitioners need to continue to educate the general public about the health benefits of healthy diets in various forms, while policy makers need to continue to promote policies that make it easier for the general public to engage in positive health behavior.

## Data Availability

The datasets analyzed during the current study are available in the Health Information National Trends Survey (HINTS) repository of the National Cancer Institute (NCI)

https://hints.cancer.gov/

## Acknowledgements

The authors wish to acknowledge Drs. Shanta Dube and Richard Rothenberg for advising JTA on her MPH thesis leading up to this manuscript

## References

1. Truglio J, Graziano M, Vedanthan R, et al: Global health and primary care: increasing burden of chronic diseases and need for integrated training. Mt Sinai J Med 79:464–74, 2012

2. Cawley J, Meyerhoefer C: The medical care costs of obesity: an instrumental variables approach. J Health Econ 31:219–30, 2012

3. Johnson NB, Hayes LD, Brown K, et al: CDC National Health Report: leading causes of morbidity and mortality and associated behavioral risk and protective factors--United States, 2005-2013. MMWR Surveill Summ 63 Suppl 4:3–27, 2014

4. Wing RR, Goldstein MG, Acton KJ, et al: Behavioral science research in diabetes: lifestyle changes related to obesity, eating behavior, and physical activity. Diabetes Care 24:117–23, 2001

5. Han JW: Physiological Index, Weight Control Behavior, and Quality of Life Related to Obesity-related Metabolic Syndrome in Korean Adults. Iran J Public Health 46:417–419, 2017

6. Burkert NT, Freidl W, Muckenhuber J, et al: Self-perceived health, quality of life, and health-related behavior in obesity: is social status a mediator? Wien Klin Wochenschr 124:271–5, 2012

7. Winslow E, Bohannon N, Brunton SA, et al: Lifestyle modification: weight control, exercise, and smoking cessation. Am J Med 101:4A25S-31S; discussion 31S-33S, 1996

8. Khan LK, Sobush K, Keener D, et al: Recommended community strategies and measurements to prevent obesity in the United States. MMWR Recomm Rep 58:1–26, 2009

9. Tsai J, Gu X: Homelessness among immigrants in the United States: rates, correlates, and differences compared with native-born adults. Public Health 168:107–116, 2019

10. Corlin L, Woodin M, Thanikachalam M, et al: Evidence for the healthy immigrant effect in older Chinese immigrants: a cross-sectional study. BMC Public Health 14:603, 2014

11. Gushulak B: Healthier on arrival? Further insight into the “healthy immigrant effect”. CMAJ 176:1439–40, 2007

12. Flores G, Brotanek J: The healthy immigrant effect: a greater understanding might help us improve the health of all children. Arch Pediatr Adolesc Med 159:295–7, 2005

13. Domnich A PD, Gasparini R, Amicizia D: The “healthy immigrant” effect: does it exist in Europe today? Ital J Public Health 9:7532, 2012

14. Agaku IT, Adisa AO, Ayo-Yusuf OA, et al: Concern about security and privacy, and perceived control over collection and use of health information are related to withholding of health information from healthcare providers. J Am Med Inform Assoc 21:374–8, 2014

15. Nelson DE, Kreps GL, Hesse BW, et al: The Health Information National Trends Survey (HINTS): development, design, and dissemination. J Health Commun 9:443-60; discussion 81-4, 2004

16. Camarota SA: Immigrants in the United States: A Profile of America’s Foreign-Born Population. Washington, DC 2012

17. Kushi LH, Byers T, Doyle C, et al: American Cancer Society Guidelines on Nutrition and Physical Activity for cancer prevention: reducing the risk of cancer with healthy food choices and physical activity. CA Cancer J Clin 56:254-81; quiz 313-4, 2006

18. Johnson RK, Appel LJ, Brands M, et al: Dietary sugars intake and cardiovascular health: a scientific statement from the American Heart Association. Circulation 120:1011–20, 2009

